# Helmet noninvasive ventilation for COVID-19 patients “Helmet-COVID”: study protocol for a multicenter randomized controlled trial

**DOI:** 10.1101/2021.08.04.21260420

**Authors:** Yaseen Arabi, Haytham Tlayjeh, Sara Aldekhyl, Hasan M Al-Dorzi, Sheryl Ann Abdukahil, Mohammad Khulaif Al Harbi, Husain Al Haji, Mohammed Al Mutairi, Omar Al Zumai, Eman Al Qasim, Wedyan Al Wehaibi, Saad Al Qahtani, Fahad Al-Hameed, Jamal Chalabi, Mohammed Alshahrani, Abdulrahman Alharthy, Ahmed Mady, Abdulhadi Bin Eshaq, Ali Al Bshabshe, Zohair Al Aseri, Zainab Al Duhailib, Ayman Kharaba, Rakan Alqahtani, Adnan Al Ghamdi, Ali Altalag, Khalid Alghamdi, Mohammed Almaani, Haifa Algethamy, Ahmad Al Aqeily, Faisal Al Baseet, Hashem Al Samannoudi, Mohammed Al Obaidi, Yassin Ismaiel, Abdulrahman A Al-Fares

## Abstract

**Introduction:** Noninvasive ventilation delivered by helmet is has been used for respiratory support of patients with acute hypoxemic respiratory failure due to COVID-19 pneumonia. The aim of this study is to compare helmet noninvasive ventilation with usual care versus usual care alone to reduce the mortality.

**Methods and analysis:** This is a multicenter, pragmatic, parallel, randomized controlled trial that compares helmet noninvasive ventilation with usual care to usual care alone in 1:1 ratio. A total of 320 patients will be enrolled in this study. The primary outcome is 28-day all-cause mortality. The primary outcome will be compared between the two study groups in the intention-to-treat and per-protocol cohorts. An interim analysis will be conducted for both safety and effectiveness.

**Ethics and dissemination:** Approvals are obtained from the Institutional Review Boards (IRBs) of each participating institution. Our findings will be published in peer-review journals and presented at relevant conferences and meetings.

**Trial registration number:** NCT04477668 registered on July 20, 2020

**Article Summary:** *Strengths and limitations of this study:*

- This trial compares helmet NIV to usual care for respiratory support of patients with acute hypoxemic respiratory failure due to COVID-19 pneumonia.
- The trial is a multi-center, pragmatic, parallel randomized controlled trial.
- The main limitation is the unblinded design due to the nature of the intervention.

## Introduction

The novel severe acute respiratory syndrome coronavirus 2 (SARS-CoV-2) has led to a pandemic resulting in over 181 million cases and approximately 3.9 million fatalities as of June 29, 2021.^1^ The resulting Coronavirus Disease 2019 (COVID-19) leads to severe pneumonia and acute respiratory distress syndrome (ARDS) among other organ injury.^2^ ARDS may occur in up to 5% of infected patients.^3-5^ Early in the pandemic, invasive mechanical ventilation was widely used because of concerns about noninvasive ventilation safety and efficacy. However, noninvasive ventilation (NIV) use increased with time, including mask NIV and helmet NIV.

NIV has been shown to be physiologic benefits in patients with acute hypoxemic respiratory failure (AHRF) secondary to pulmonary edema, atelectasis, or pneumonia.^6^ It has been shown to improve arterial oxygenation by increasing functional residual capacity, shifting the tidal volume to a more compliant part of the pressure-volume curve, thus reducing both the work of breathing and the risk of tidal opening and closure of the airways.^7^ NIV is commonly provided through nasal or oro-nasal interfaces. The resulting aerosol generation may increase the risk of transmission of pathogens to healthcare providers, raising concerns about the use of NIV in patients with viral pneumonia.^8^ Helmet NIV has been used for AHRF including patients with COVID-19 pneumonia. The helmet surrounds the patient’s entire head to provide positive pressure and supply oxygen and is sealed with a soft, airtight collar that wraps around the neck. Due to this design, it has advantages over the nasal and oro-nasal interfaces. These include less air leaks, no skin or nasal bridge skin injuries, no eye irritation, fitting for different facial contours,^9^ and hypothetically less dissemination of aerosols in the environment. However, helmet interface may be associated with increase in dead space (especially if the settings are not used appropriately), claustrophobia, discomfort, and difficulty in access for suction and feeding.

Evaluation of helmet NIV as a respiratory support modality started more than two decades ago.^10^ Helmet NIV has been investigated as a treatment for different forms of AHRF in adults in various settings, such as pre-hospital ambulance, emergency department and ICU.^7 11-15^ However, earlier clinical studies are relatively scarce and mostly small in size, and often used improvement in oxygenation and intubation rate as primary outcomes.^6^

However, the evidence on helmet NIV for AHRF is growing. A systematic review of randomized controlled trials (RCTs) and observational studies published before June 2016 found 11 studies involving 621 patients.^16^ Compared with controls, the use of the helmet was associated with lower hospital mortality, intubation rate, and complications.^16^ Moreover, there was no significant difference in gas exchange and ICU stay.^16^ A meta-analysis of four RCTs (377 patients) showed that helmet NIV significantly increased the ratio of arterial oxygen partial pressure to fraction of inspired oxygen (PaO_2_/FiO_2_) and decreased arterial carbon dioxide levels, intubation rate and in-hospital mortality compared to standard oxygen therapy.^17^ In a more recent systematic review and network meta-analysis that included 25 studies (published up to April 2020) and 3804 patients with AHRF, mortality and intubation rate were lower with helmet NIV compared to standard oxygen by more than 50%, while the effect of mask NIV and high-flow nasal oxygen were modest compared to standard oxygen.^18^ Helmet NIV was superior to both mask NIV and high-flow nasal oxygen, while mask NIV and high-flow nasal oxygen were not different in their effects on mortality and intubation rate.^18^ One study reported the cost-effectiveness of helmet NIV compared to mask NIV.^19^

Data on helmet NIV in AHRF related to COVID-19 are emerging. A recent RCT conducted in 4 Italian ICUs on patients with COVID-19 and moderate to severe AHRF found that treatment with helmet NIV did not result in significantly fewer days of respiratory support at 28 days from randomization (primary outcome) as compared with high-flow nasal oxygen alone (mean difference 2 days, 95% CI, -2 to 6, p=0.26).^20^ Nevertheless, the intubation rate was significantly lower in the helmet NIV group compared to the high-flow nasal oxygen group (30% vs 51%; p=0.03).^20^ Additionally, the median number of days free of invasive mechanical ventilation within 28 days was significantly higher in the helmet NIV group than in the high-flow nasal oxygen group (28 versus 25 days; mean difference, 3 days; 95% CI, 0-7; P□= □0.04).^20^ The hospital mortality was 24% in the helmet NIV group and 25% in the high-flow nasal oxygen group.^20^

As the efficacy of helmet NIV to improve outcomes in severe AHRF due to COVID-19 pneumonia has not been clearly established, the aim of this study is to compare helmet NIV with usual care versus usual care alone to reduce 28-day all-cause mortality. We hypothesize that helmet NIV will reduce 28-day all-cause mortality in patients with suspected or confirmed severe COVID-19 pneumonia and AHRF.

## Methods and analysis

### Trial design

This is an investigator-initiated, pragmatic parallel RCT that will compare helmet NIV with usual care to usual care alone in 1:1 ratio in patients with suspected or confirmed COVID-19 pneumonia and AHRF. Randomization is performed using a computer-generated schedule using variable block sizes (4 or 6) and is stratified by site. The trial is sponsored by King Abdullah International Medical Research Center, Riyadh, Saudi Arabia, has been registered with ClinicalTrials.gov (NCT04477668) and is conducted across multiple centers in Saudi Arabia. Training and in-service education on helmet NIV use as well as on protocol implementation are provided to all participating centers. The competency of the bedside respiratory therapists is supervised by experienced respiratory care supervisors and intensivists.

### Sample size

In a large observation study of patients with AHRF and ARDS, hospital mortality was 34.9% (95% CI, 31.4-38.5%) for patients with mild ARDS, 40.3% (95% CI, 37.4%-43.3%) for those with moderate ARDS, and 46.1% (95% CI, 41.9%-50.4%) for those with severe ARDS.^21^ A systematic review found an overall pooled mortality estimate among 10,815 patients with ARDS due to COVID-19 to be 39% (95% CI: 23–56%).^22^ Considering a mortality rate of 40% in patients with COVID-19 pneumonia and moderate to severe ARDS treated with usual care, we calculated that enrollment of 304 patients (152 in each group) would provide the study with 80% power to demonstrate an absolute difference of 15% in the primary outcome between the usual care group and helmet NIV group at a two-sided alpha level of 0.05. To account for 5% loss to follow-up, the total number of patients required for the trial is 320 patients.

### Participant eligibility

The trial will enroll ICU patients with suspected^24^ or confirmed COVID-19 pneumonia who have AHRF. Detailed inclusion and exclusion criteria can be found in **Table 1**.

**Table 1.**
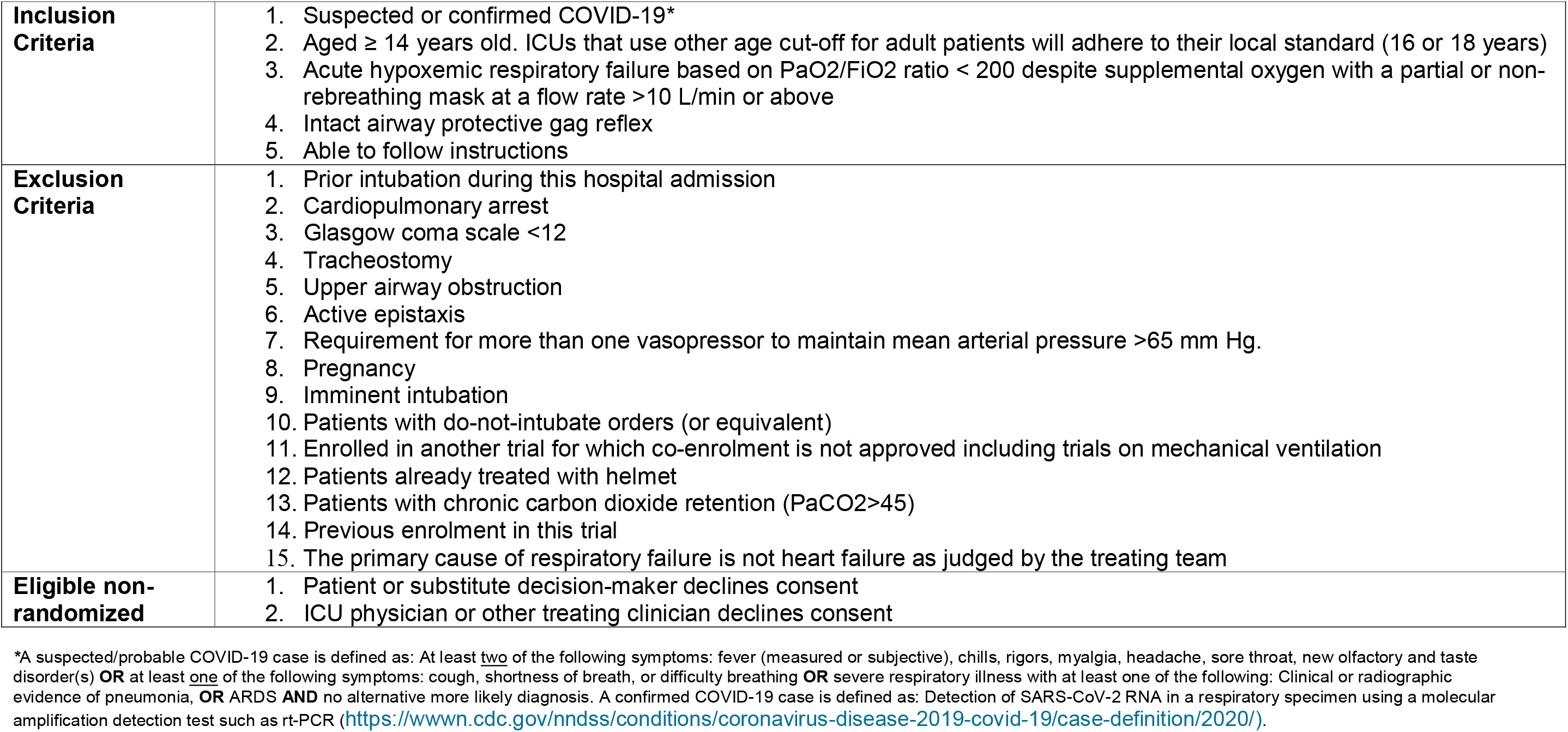
Eligibility Criteria

|  |  |
| --- | --- |
| **Inclusion Criteria** | 1. Suspected or confirmed COVID-19*<br>2. Aged $\geq 14$ years old. ICUs that use other age cut-off for adult patients will adhere to their local standard (16 or 18 years)<br>3. Acute hypoxemic respiratory failure based on PaO<sub>2</sub>/FiO<sub>2</sub> ratio $< 200$ despite supplemental oxygen with a partial or non-rebreathing mask at a flow rate $>10$ L/min or above<br>4. Intact airway protective gag reflex<br>5. Able to follow instructions |
| **Exclusion Criteria** | 1. Prior intubation during this hospital admission<br>2. Cardiopulmonary arrest<br>3. Glasgow coma scale $<12$<br>4. Tracheostomy<br>5. Upper airway obstruction<br>6. Active epistaxis<br>7. Requirement for more than one vasopressor to maintain mean arterial pressure $>65$ mm Hg.<br>8. Pregnancy<br>9. Imminent intubation<br>10. Patients with do-not-intubate orders (or equivalent)<br>11. Enrolled in another trial for which co-enrolment is not approved including trials on mechanical ventilation<br>12. Patients already treated with helmet<br>13. Patients with chronic carbon dioxide retention (PaCO<sub>2</sub>$>45$)<br>14. Previous enrolment in this trial<br>15. The primary cause of respiratory failure is not heart failure as judged by the treating team |
| **Eligible non-randomized** | 1. Patient or substitute decision-maker declines consent<br>2. ICU physician or other treating clinician declines consent |

\*A suspected/probable COVID-19 case is defined as: At least two of the following symptoms: fever (measured or subjective), chills, rigors, myalgia, headache, sore throat, new olfactory and taste disorder(s) **OR** at least one of the following symptoms: cough, shortness of breath, or difficulty breathing **OR** severe respiratory illness with at least one of the following: Clinical or radiographic evidence of pneumonia, **OR** ARDS **AND** no alternative more likely diagnosis. A confirmed COVID-19 case is defined as: Detection of SARS-CoV-2 RNA in a respiratory specimen using a molecular amplification detection test such as rt-PCR (https://wwwn.cdc.gov/hndss/conditions/coronavirus-disease-2019-covid-19/case-definition/2020/).

### Informed consent

Informed consent will be obtained from the potential trial participants or their surrogate decision makers. A hybrid model of consent will be used where a priori consent is obtained if possible, otherwise delayed consent model will be obtained as per local approvals. The first patient was enrolled in February 2021. As of June 29, 2021, a total of 199 patients were enrolled from 5 sites. There are several sites that are processing IRB and regulatory approvals.

### Patient and public involvement

There was no patient or public involvement in the conception, design or conduct of the study, or the writing or editing of this paper. However, patient comfort and experience as well as compliance to the intervention were taken in to consideration and data on these are collected.

### Trial interventions

#### Helmet group

A helmet (Subsalve, USA or its equivalent), which is made of transparent latex-free polyvinyl chloride, will be applied to patients randomized to the intervention group as per the study protocol which considers the manufacturer instructions. It will be immediately connected to an ICU ventilator in pressure support (PS) mode with positive end-expiratory pressure (PEEP) using a conventional respiratory circuit joining two port sites to allow inspiratory and expiratory flow. The starting settings is PS of 8-10 cm H2O, PEEP of 10 cmH2O with FiO_2_ of 100%, targeting flow rate of ≥50 L/min with an inspiratory rise time of 50 msec and end flow/cycling off of 50% of maximal inspiratory flow. PEEP may be increased by 2 cm every 3 minutes to achieve oxygen saturation (SpO2) ≥90% on FIO_2_ ≤ 60%, and PS can be increased by 2 cm every 3 minutes to achieve respiratory rate ≤ 25/min and disappearance of accessory muscle activity. The maximal allowed PS + PEEP is 30 cm H_2_O. Interruptions of helmet should be avoided or kept at minimum at least in the first 48 hours.^20^ More details of helmet NIV application, set up and weaning can be found in the **Supplementary File**. Some patients may not tolerate helmet NIV. In that case, the physician or the respiratory therapist explains the procedure to the patient. Dexmedetomidine infusion may be used to improve ccomfort with the helmet NIV. Other intravenous sedatives such as benzodiazepines or intravenous narcotics should generally not be used. If the patient continues to be intolerant to the helmet, the patient can be managed according to the usual care. Detailed data about helmet NIV tolerance are collected.

#### Control group

In the control group, patients receive usual care according to the clinical practices of each site. This may include oxygen provided using standard oxygen devices, high-flow nasal oxygen or NIV provided by nasal mask, face mask or total mask.

### Endotracheal Intubation

The decision to intubate will be at the discretion of the treating team with no involvement from the research team. However, the protocol provides guidance on assessing patients for the need of endotracheal intubation throughout the study period (for both study groups: helmet NIV or usual care) according to the following general principles:

Enrolled patients should be assessed within 4 hours of enrollment (or sooner as required) and at frequent intervals for the following criteria, although the decision is usually not based on a single variable:

- Neurologic deterioration *(not attributed to sedation)*
- Persistent or worsening respiratory failure of NIV (manifesting as oxygen saturation <88%, respiratory rate >36/min, PaO_2_/FiO_2_ ratio <100 or persistent requirement of FiO_2_ ≥70%)
- Intolerance of face mask or helmet
- Airway bleeding
- Copious respiratory secretions
- Respiratory acidosis with pH <7.25
- Hemodynamic instability
- Significant radiologic worsening

### Co-Interventions

Patients who require endotracheal intubation are managed by the primary team with lung protective strategy with tidal volumes of 6 mL/kg of predicted body weight and titration of PEEP to achieve oxygen saturation of 88% to 95% at the lowest possible FiO_2_. Daily interruption of sedation, awakening and breathing trials, and early mobilization are performed as per the ICU standards.^25^ Management of COVID-19 is provided as per local protocols; physicians are advised to follow the clinical practice guidelines set by the Saudi Critical Care Society,^26^ the Surviving Sepsis Campaign,^27 28^ and the World Health Organization.^29^ The study protocol does not mandate particular therapies; however, corticosteroids, immune modulators and antiviral therapy are all recorded. Conservative fluid management is recommended where neutral balance should be targeted and intravenous resuscitation should be reserved for shock treatment in both groups and fluid balance is recorded.

### Blinding

Due to the nature of the study intervention, blinding is not be possible.

### Recruitment schedule and enrollment procedures

Schedule of assessments is detailed in **Table 2**. All non-intubated subjects with suspected or confirmed COVID-19 are screened upon admission to the ICU. A screening log will be kept to monitor and report the size of the patient population from which eligible patients have been randomized. Co-enrollment in other RCTs is permissible as long as inclusion in the other RCT would not confound the results of this trial and after discussion with the steering committees of the other studies.

**Table 2.** Schedule of assessments in the trial

| **Task** | **Screening** | **Randomization** | **Baseline** | **Day 1-28** | **180-Day**<br>**Follow up** |
| --- | --- | --- | --- | --- | --- |
| Assess eligibility to enter study | X |  |  |  |  |
| Assess ability to gain consent & follow-up | X |  |  |  |  |
| Consent | X |  |  |  |  |
| Demographics and eligibility checklist | X | X | X |  |  |
| Laboratory data |  |  | X | X |  |
| Vital signs |  |  | X | X |  |
| Vital status up to Day 28 in the ICU |  |  |  | X | X |
| Vital and functional status |  |  |  |  | X |
| Discharge date from ICU, from hospital |  |  |  | X | X |
| Adverse events |  |  |  | X |  |
| Protocol violations |  |  |  | X |  |

### Data collection

Baseline data on demographics, admission diagnosis and clinical information are collected. Clinical information include Acute Physiology and Chronic Health Evaluation (APACHE) II score,^30^ source of admission, ICU admission category (elective, emergency or non-surgical), ICU admission diagnosis and co-morbidities (as defined by the APACHE II severity of illness scoring system). Daily data will be recorded until discharge from ICU or 28 days after randomization. We will collect data on the use of helmet including the tolerance of helmet (>1-hour use).

### Outcomes

The primary outcome is 28-day all-cause mortality. Secondary outcomes are intubation rate within 28 days, ICU mortality, hospital mortality (censored at day 180), ICU-free days at day 28, invasive ventilation-free days at day 28, renal replacement therapy-free days at day 28 and vasopressor-free days at day 28. Safety outcomes include skin pressure injuries, barotrauma and serious adverse events (including cardiovascular events and device complications).

Additionally, there will be a follow up of enrolled patients at day 180 about vital status, functional status (EuroQoL (EQ)-5D-5L) which is planned to be reported separately. For patients who have been discharged from hospital before day 180, follow up will be conducted by telephone.

### Data analysis

A formal statistical analysis plan will be agreed upon and placed in the public domain before the study database is locked for the analysis of the primary outcome. The primary outcome will be compared in the intention-to-treat and per-protocol cohorts (effectiveness analysis) using the chi-square test. Results will be reported as relative risk with 95% CI. Kaplan–Meier curves will be plotted to assess the time from enrollment to death and will be compared by means of the log-rank test. A two-tailed P value <0.05 will be considered to indicate statistical significance. SAS software, version 9.2 (SAS Institute) will be used for all the analyses.

A priori analysis will be done for the following subgroups:

I. Patients with moderate ARDS (PaO_2_/FIO_2_ ratio 100-200) and patients with severe ARDS (PaO_2_/FIO2 ratio <100)
II. Obese patients (body mass index >30 kg/m^2^) and patients with body mass index ≤30
III. Patients aged >65 years and ≤65 years
IV. APACHE II score higher or lower than the median of enrolled patients

For the occasional randomized patient who is withdrawn from the trial and allows use of data, the patient’s data will be included in the group to which he/she was allocated as per the intention-to-treat principle and the reason of withdrawal will be documented.

### Trial management and monitoring

The study Steering Committee members will be responsible for overseeing the conduct of the trial, for upholding or modifying study procedures as needed, addressing challenges with protocol implementation, formulating the analysis plan, reviewing and interpreting the data, and preparing the manuscript. This will be achieved through meetings (in-person or by conference calls) at least quarterly.

Several measures are taken to minimize, observe and document any potential safety concerns. First, any unexpected safety concerns will be reported immediately to the Steering Committee and IRB. Second, an independent Data Safety Monitoring Board will be monitoring the safety of the trial. Lastly, interim analyses will be conducted after recruiting 1/3 and 2/3 of the total patients and the interim test statistics will be the primary outcome analysis for both safety and effectiveness. The Data Safety Monitoring Board will use formal stopping rules based on the primary endpoint of 28-day mortality. The trial may be stopped for safety (p<0.01) or effectiveness (p<0.001). There will be no plans to terminate the trial for futility. We will account for alpha spending by the O’Brien Fleming method and the final p value will be considered at 0.048. The principles used in the conduct of safety monitoring and reporting in this trial are those outlined by Cook et al.^32^

In this trial, reporting of serious adverse events will be restricted to events that are not captured as study outcome and are considered to be related to the helmet NIV (possibly, probably or definitely).^32^ These may include cardiovascular events (i.e., cardiac arrest and hypotension with drop in blood pressure to systolic <90 mm Hg) and device complications (i.e., helmet deflation).

### Ethics and dissemination

The study will be conducted according to the principles of the latest version of Good Clinical Practice (GCP) and in accordance with all relevant local ethical, regulatory, and legal requirements. A manuscript with the results of the primary study will be published in a peer-reviewed journal. Separate manuscripts will be written on secondary aims, and these will also be submitted for publication in peer reviewed journals as well.

## Discussion

The importance of this study stems from the current pandemic situation as different treatment modalities are being sought to answer important clinical questions. Available literature on the evaluation of helmet NIV as a respiratory support modality in COVID-19 patients is limited. Table 3 provides the list of ongoing RCTs on helmet NIV. This study aims to contribute to the existing literature and in turn influence clinical practice.

**Table 3.** List of ongoing registered randomized controlled trials (RCTs) on helmet noninvasive ventilation

| **Trial** | **Registration** | **Interventions** | **Design** | **Countries** | **N** |
| --- | --- | --- | --- | --- | --- |
| Helmet non-invasive ventilation for COVID-19 patients (Helmet-COVID) | NCT04477668 | Helmet vs. usual care | Multicenter RCT | Saudi Arabia | 320 |
| Comparison of high-flow nasal oxygen, face-mask NIV and helmet NIV in COVID-19 ARDS patients (NIV COVID19) | NCT04715243 | High-flow nasal oxygen vs. helmet NIV vs. mask NIV | Multicenter RCT | Oman | 360 |
| Helmet CPAP vs. high-flow nasal oxygen in COVID-19 (COVID HELMET) | NCT04395807 | High-flow nasal oxygen vs. helmet CPAP | Single-center RCT | Sweden | 120 |
| high-flow nasal oxygen vs. CPAP Helmet in COVID-19 pneumonia (COVIDNOCHE) | NCT04381923 | High-flow nasal oxygen vs. helmet CPAP | Single-center RCT | USA | 200 |
| Early CPAP in COVID-19 patients with respiratory failure (EC-COVID-RCT) | NCT04326075 | Early helmet CPAP vs. usual care | Single-center RCT | Italy | 900 |

We planned our pragmatic trial to address whether using helmet NIV as the primary non-invasive respiratory support in patients with severe COVID-19, in addition to the commonly used high-flow nasal oxygen and mask NIV improves outcome. By nature of this question, there is heterogeneity of the control group; as patients in this group could receive standard oxygen, high-flow nasal oxygen or mask NIV at the decision of the treating team. This approach is supported by a recent network meta-analysis of randomized controlled trials that showed only a modest effect of high-flow nasal oxygen and mask NIV on mortality or intubation rate compared to standard oxygen, while patients treated with helmet NIV had more than 50% reduction in mortality and intubation rate compared to the other three modalities.^18^ In addition, this approach is likely to be more representative of usual practice in which patients may get oxygen therapy, high-flow nasal oxygen and NIV at different times during their acute illness. Given the fact that the use of helmet NIV has not been widespread across ICUs, we thought that the broader question addressed by our study might be more relevant to deciding whether to introduce this modality or not in a given ICU.

The main limitation to our study is inability to blind the given allocation due to the nature of the intervention.

## Data Availability

Data will be provided upon reasonable request from the Chief Investigator.

## Acknowledgements

The authors would like to thank all the participating patients and their families, as well as the members of the Data Safety & Monitoring Board: Chair: Dr. Nicholas S. Hill (Professor of Medicine, Chief of Pulmonary, Critical Care and Sleep Division, Tufts Medical Center, Boston, Massachusetts, USA), DSM members: Dr. Stefano Nava (Professor of Respiratory Medicine University of Bologna, Chief of the Respiratory and Critical Care Sant’ Orsola Hospital Bologna, Specialist in Respiratory Medicine and Intensive Care Medicine, University of Bologna, Italy), Dr. James Mojica (Vice Chief and Clinical Director of Pulmonary & Critical Care, Director, The Sleep Center at Spaulding, Massachusetts General Hospital, USA) and Dr. Michael Harhay (Assistant Professor of Epidemiology and Medicine-Pulmonary and Critical Care, Department of Biostatistics, Epidemiology and Informatics, University of Pennsylvania USA).

## Contributors

YA is the principal investigator and participated in the project concept, design, final approval, and manuscript preparation, review and submission. HT, SD, HD, SA, MH, HH, MM, OZ, EQ, WW, SQ, FH, JC, MS, AH, AM, AE, AB, ZA, ZD, AK, RQ, AG, AT, KG, MA, HG, AA, FB, HS, MO, YI, AF participated in the critical revision, final approval of the protocol and manuscript preparation, review and submission. All authors agree to be accountable for the accuracy and integrity of the work.

## Funding

The study is funded by King Abdullah International Medical Research Center (RC 20/306/R). The study sponsor does not have any role in the study design, collection, management, analysis, and interpretation of data as well as writing of the report.

## Competing interests

None declared

## Ethics approval

The study has been approved by the Institutional Review Board (IRB) of Ministry of National Guard Health Affairs (MNGHA) and the respective Institutional Review Boards of all the other centers.

## Patient consent for publication

Not required.

## Provenance and peer review

Not commissioned, externally peer reviewed.

## Notes

### Competing Interest Statement

The authors have declared no competing interest.

### Clinical Trial

NCT04477668

### Author Declarations

The study has been approved by the Institutional Review Board (IRB) of Ministry of National Guard Health Affairs (MNGHA) and the respective Institutional Review Boards of Imam Abdulrahman bin Faisal University; King Saud Medical City ;King Khalid Hospital, Najran; Aseer Central Hospital; King Faisal Specialist Hospital & Research Center, Riyadh; King Fahad Hospital Madinah; King Khalid University Hospital, Riyadh; Prince Sultan Military Medical City; King Abdulaziz University Hospital Jeddah and Al Amiri Hospital Kuwait

