## Supplementary file for "Helmet noninvasive ventilation for COVID-19 patients “Helmet-COVID”: study protocol for a multicenter randomized controlled trial"

**Helmet-NIV Protocol**

**Setup and preparation**

- Use unheated, low-compliance, dual limb breathing circuit.
- Insert a bacterial/viral filter at the expiratory port of the ventilator.
- Carry out the required preoperational check.
- To avoid patient-ventilator asynchrony, explain the application and the procedure of the helmet to the patient.
- Insert the earplugs into patient ears.
- Use the under-arm pads to avoid strap pressure.

**Initial setup**

- Can be used with a ventilator or NIV machines.
- Set up the ventilator on Pressure Support Ventilation (PSV).
- PSV 8-10 cm H2O pressure, positive end-expiratory pressure (PEEP) 10 cm H2O pressure, FiO2 100%.
- Flow rate >50 L/min, inspiratory rise time 50 msec, End Flow/Cycling off 50% of maximal inspiratory flow.

**Titration**

- Increase PEEP by 2 cm H2O every 3 minutes to achieve SpO2 ≥ 92% on FiO2 ≤ 60%
- Higher PEEP than what is used on face mask NIV is allowed and tolerated.
- Increase PS by 2 cm every 3 minutes to achieve respiratory rate (RR) ≤ 25/min or clear patient comfort.
- The maximal allowed PS + PEEP is 30 cm H_2_O.
- Helmet interruptions should be avoided or kept at minimum at least for the first 48 hours.
- Titrate FiO2 to ≤60% as soon as possible.

**Sedation**

- Dexmedetomidine may be used to improve compliance with the helmet. Other intravenous sedatives such as benzodiazepines or intravenous narcotics should not be used.

**Weaning when clinical condition allows**

- Titrate PS by 2 cm H2O every 3 hours if RR ≤ 25/min.
- Titrate PEEP by 2 cm H2O every 3 hours if SpO2 >92% on <60% FiO2.
- If RR ≤25/min on PSV ≤8 cm and SpO2 >92% on FiO2 ≤50% and PEEP ≤8 cm H2O, the helmet can be discontinued and patient switched to high-flow nasal cannula, or oxygen supply at O2 at 6L/min or higher.
- Helmet NIV could be resumed at any time if the respiratory rate was greater than 25 breaths/min and/or SpO2 was lower than 92% on FiO2 of ≥60%.

**Nursing Care**

- Perform oral care/suction before helmet application.
- Nutrition can be provided through a straw. Place a nasogastric tube before helmet application if felt necessary by treating physician (not commonly needed).
- A suction Yankauer for suctioning and a straw for drinking can be introduced through the patient port without removing the helmet.
- Intravenous sedation (Dexmedetomidine or others) can be used at the discretion of the treating physician to assure patient comfort.

**Equipment required**

- Mechanical ventilator (BP 840, PB 980, Servo-I, Drager V 500, Astral and V680)
- Unheated double lumen circuit
- Bacterial/viral filter
- Subsalve helmet or equivalent
- Under arm pads
- Ear plugs

**General recommendations to consider intubation for patients on NIV (assessed within 4 hours and at frequent intervals throughout NIV treatment):**

- Neurologic deterioration (not attributed to sedation)
- Persistent or worsening respiratory failure of NIV:
  - oxygen saturation <88%
  - respiratory rate >36/min
  - P/F ratio <100
  - a persistent requirement of FiO2 ≥70%
- Intolerance of face mask or helmet
- Airway bleeding
- Copious respiratory secretions
- Respiratory acidosis with pH <7.25
- Hemodynamic instability
- Significant radiologic worsening

**Humidification**

Appropriate level of humidification can be achieved via bubble humidifier with external oxygen flow of 5 L/Min entrained into the ventilator circuit proximal to the patient helmet.
